# Machine Learning Assisted Differentiation of Low Acuity Patients at Dispatch (MADLAD): A Randomized Controlled Trial

**DOI:** 10.1101/2025.09.19.25336143

**Authors:** Douglas Spangler, Simon Morelli, David Smekal, Lennart Edmark, Hans Blomberg

## Abstract

**Background:** Resource Constrained Situations (RCS) at Emergency Medical Dispatch centers where there are more patients requiring an ambulance than there are available ambulances are common. Machine learning (ML) techniques offer a promising but largely untested approach to assessing relative risks among these patients. The study aims to establish whether the provision of ML-based risk scores predicting patient outcomes improves the ability of dispatchers to identify patients at high risk for deterioration in RCS and dispatch the first available ambulance to them.

**Methods:** A parallel-grouped, randomized trial of adult patients assessed by a dispatch nurse in the Swedish regions of Uppsala or Västmanland as requiring a low-priority ambulance response in RCS. Patients were randomized 1:1 to be prioritized with the aid of a ML-based risk assessment tool, or per current clinical practice. Prioritization accuracy was assessed primarily in terms of whether the first available ambulance was sent to the patient with the highest National Early Warning Score (NEWS 2) based on subsequently collected vital signs. Trial registered at ClinicalTrials.gov (NCT04757194).

**Results:** A total of 1245 RCS were included in the study. In the intervention group, patients assigned the first available ambulance had the highest NEWS in 68.3% of cases vs 62.5% in the control group, corresponding to an odds ratio of 1.28 (95% CI 1.00 – 1.63, p = 0.047). Prespecified analyses also suggested that dispatchers complied with the tool in 80.9% (77.7 – 83.9) of cases, and that full compliance with the risk prediction instrument would have improved prioritization decisions further.

**Discussion:** This study suggests that clinical ML-based decision support tools have the ability to influence care provider decisions and improve their capacity to rapidly differentiate between high- and low-risk patients at dispatch.

## Introduction

In prehospital care systems, ambulance availability places constraints on the ability of Emergency Medical Dispatch (EMD) centers to immediately provide an ambulance response for all patients determined to require one. The stochastic nature of ambulance demand via the emergency hotline entails that any cost-effective ambulance system will from time to time experience Resource Constrained Situations (RCS) in which the number of patients requiring an ambulance response exceeds the number of available ambulances.

Machine learning (ML) models offer a promising approach to stratifying the risks associated with these patients. In previous research, an open source ML-based risk assessment tool was developed and retrospectively validated in a cohort of patients receiving ambulance care in the region of Uppsala (1,2). While a number of ML-based risk assessment tools have been proposed for use in prehospital- and emergency care and validated retrospectively (3–9), only a single randomized trial has been performed (10,11) and no models intended for risk differentiation across the full spectrum of prehospital patient types have been evaluated in a randomized trial. There is thus a great need to identify suitable use-cases for these tools, and to generate high-quality evidence regarding their effectiveness in achieving clinically important objectives. Given that the application of ML models in this context is novel and unproven, this must be done in a manner which minimizes the patient safety hazards associated with incorrect decisions.

### Aim

This study aims to investigate whether the application of a ML-based risk assessment instrument improves the ability of dispatchers to identify and dispatch an ambulance to the most critically ill patient in RCS. Patient acuity was operationalized primarily as the National Early Warning Score (NEWS 2 base scale) based on the first set of vital signs obtained, and secondarily based on a composite risk score consisting of prehospital interventions and hospital outcomes.

### Hypotheses

#### Primary

1. The intervention results in a greater proportion of immediate ambulance responses in RCS being directed to the patient in the most critical condition as operationalized by subsequent NEWS value.

#### Secondary

1. The intervention improves differentiation with regards to a composite risk score consisting of ambulance interventions, abnormal initial ambulance findings, emergent transport, hospital admission, and mortality between patients receiving immediate vs. delayed ambulance response during RCS.
2. The intervention increases the difference in NEWS between patients receiving immediate vs. delayed ambulance response during RCS.

#### Pre-specified ancillary analyses

1. Evaluation of overall personnel compliance with risk assessment instrument in intervention arm.
2. Evaluation of compliance in intervention arm cases where the model had a high vs low level of confidence.
3. Evaluation of improved/degraded compliance with risk assessment instrument over time as manifested by a slope change in a time series analysis of intervention group
4. Evaluation of spillover effects as manifested by a significant positive slope in a time series analysis of control group outcomes.
5. Evaluation of change in risk assessment tool predictive value over time (covariate drift).
6. Evaluation of model calibration with regards to age, gender, and complaint category.

## Methods

### Design

A parallel grouped trial, randomized 1:1 to intervention or control arms.

### Setting

The study took place in two EMDCs in central Sweden (Uppsala and Västmanland), serving a combined population of 499 000 in 2021. The regions have a total of 32 ambulances during peak hours. Each dispatch center is staffed by 2-3 dispatch nurses that answer emergency (112) and non-emergency calls, and 1 ambulance director 24 hours per day. The dispatch nurses currently employ a self-developed, rule-based Clinical Decision Support System (CDSS) to structure patient interviews and determine a priority level (12,13). The CDSS results in a priority of 1A/1B (lights and sirens response) 2A/2B (non-emergency response) or referral to non-ambulance care. Priority 2A/2B patients were chosen as the target population for the intervention, given that they had been determined by a dispatch nurse to be relatively low acuity, but would also not be exposed to the risks associated with being referred to non-emergency care (12,14,15).

### Participants

#### Inclusion Criteria

- Identification of a resource constrained situation by ambulance director (i.e., 2 or more patients awaiting an ambulance response).
- Assigned priority 2A or 2B by dispatch nurse.
- Complete call documentation in the CDSS.
- Valid Swedish personal identification number collected at dispatch.
- Age >= 18 years.

#### Exclusion Criteria

- Relevant calls received more than 30 minutes apart.
- Logistical factors (e.g. the patients’ geographical locations) affect the ambulance assignment decision.
- On scene risk factors (e.g. a patient is outdoors and risks hypothermia) or risk mitigators (e.g. healthcare staff already on-scene with a patient) affect the ambulance assignment decision.

### Intervention

Ambulance directors had overall responsibility for executing the study protocol, and were tasked with identifying RCS suitable for inclusion in the study. Patients were to be included in the study upon the identification of an RCS involving eligible patients by the ambulance director at the point in time when an ambulance was available for dispatch to one of the patients. Directors were instructed to consider any relevant non-clinical factors prior to randomization, and exclude any RCS where these factors would override a clinical determination per the above exclusion criteria. The above inclusion criteria were applied automatically (i.e., patients not meeting criteria could not be selected), while exclusion criteria were applied by ambulance directors. Upon selecting the relevant patients and pressing a button in the dispatch interface to compare the selected patients, the RCS was randomly assigned to a study arm via a random number generator. In the control arm, the risk scores for each patient were calculated and stored, but not displayed to the user. In the intervention arm, a mark was displayed in the interface indicating which of the included patients had the highest risk score, along with a color-coded indicator of model confidence (red for high or orange for low, with a cutoff value calibrated to include ca. 50% of patients in each group). Figure 1 below illustrates the user interface presented to the dispatcher, with Fig 1A illustrating the cases having been selected for inclusion but prior to the user pressing the “predict” button, and Fig 1B illustrating a comparison included in the intervention arm with a high level of model confidence in the selected high-risk patient.

**Figure 1.**
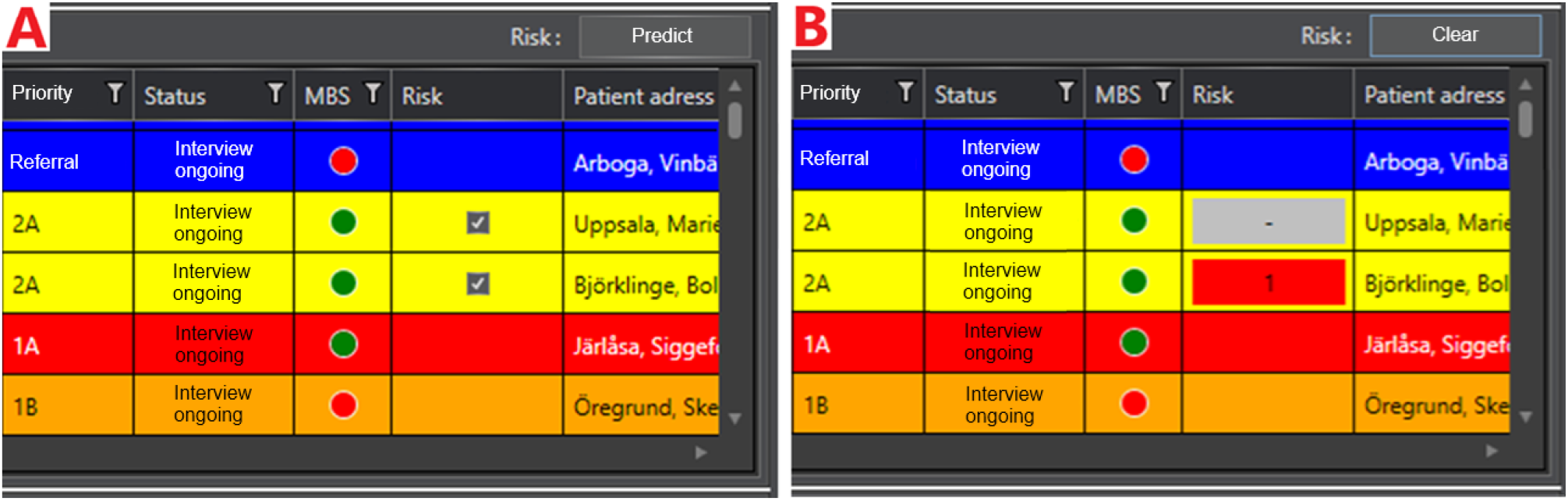
Illustration of Graphical User Interface (translated)

**Figure 2.**
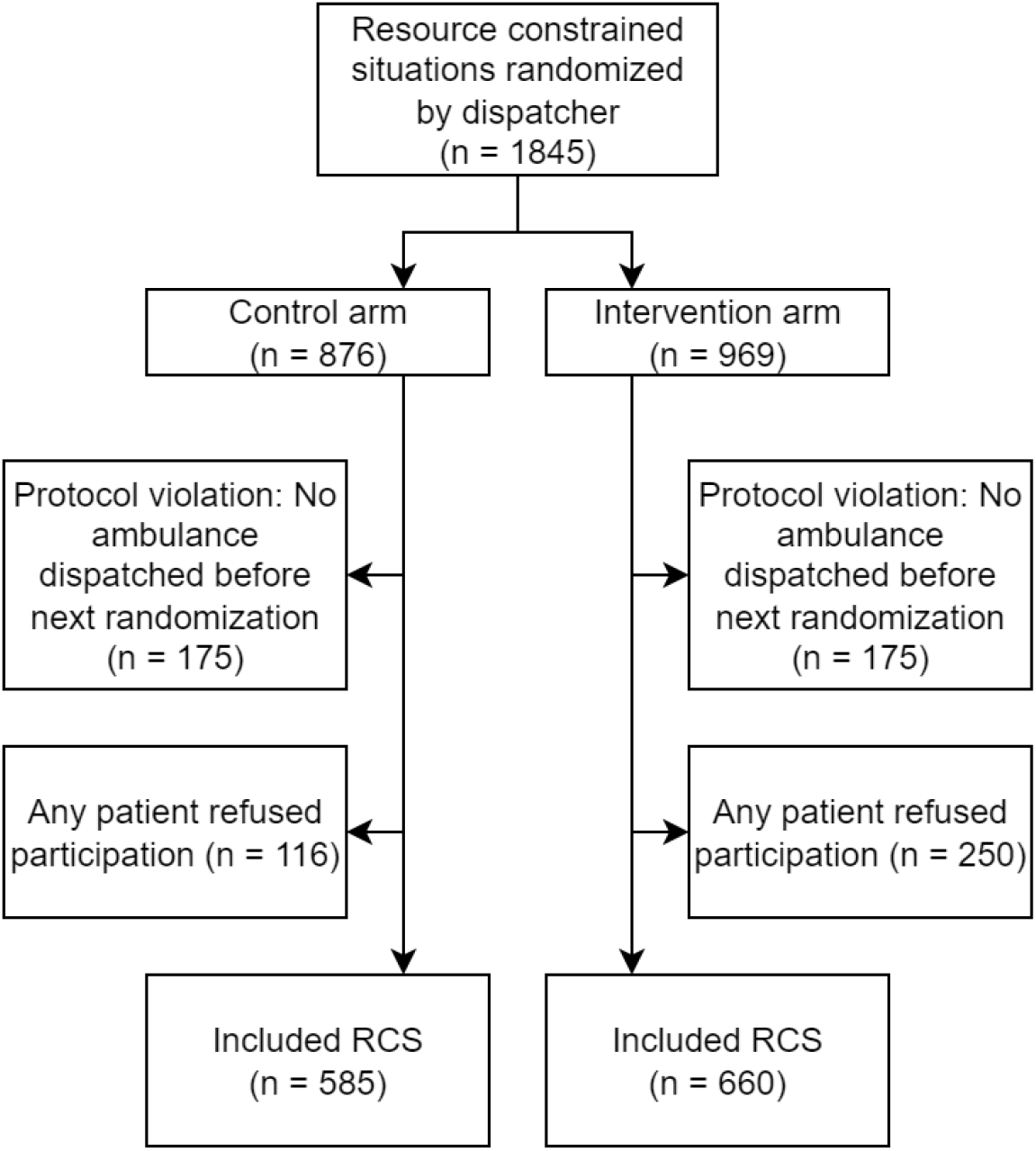
Participant flow chart

In both study arms, the ambulance director then conferred with the nurses involved in triaging the patients to confirm which patient should receive the available ambulance. In the intervention arm, the ambulance director noted which patient was proposed by the ML framework, and could access additional information regarding the risk assessment by clicking the risk buttons in the interface. The director then dispatched the available ambulance to the patient determined through this process to have the greatest need and cleared the prediction. This process was repeated each time an ambulance became available.

The intervention was based on a risk assessment instrument validated in a previous study (1). Since the publication of the validation study, the risk assessment instrument was further developed to include free-text notes entered by dispatchers, which were found to improve the performance of the models. The source code of the tool employed in the study is available on github under an open-source license (2). The tool estimates the likelihood that a patient will be assessed by ambulance crews to 1) have abnormal initial findings, 2) be transported to the hospital with lights and sirens, 3) receive a prehospital intervention, and 4) be admitted to the hospital or die within 30 days. Hospital outcome measures are based on the first hospital visit within 72 hours, in order to capture hospital outcomes for non-conveyed patients. The predicted likelihood for each of the outcomes was then combined into a composite risk score, with the above outcomes weighted to achieve predictive properties similar to those of NEWS, resulting in weights of 4:2:1:1, respectively. Development of the modelling framework was frozen upon initiation of the main study phase and was updated only upon identification of negative trends in model accuracy based on assessment by the data monitoring committee, and upon initiation of the study at the second site.

### Outcomes

The primary outcome of the study was the National Early Warning Score (NEWS) of each included patient, based on the first set of vital signs captured by the ambulance crew upon arrival to the patient. Where vital signs were not documented by an ambulance (e.g., if a patient used an alternate mode of transport to the hospital), NEWS component items were multiply imputed including the first set of vital signs documented within 24 hours from the emergency department. NEWS was selected as the primary outcome of the study for two reasons: Firstly, NEWS is widely used in acute care, and has been thoroughly validated as being predictive of outcomes in a variety of adult patient cohorts. (16–19) Secondly, NEWS is based on patient vital signs, and is thus conceptually distinct from-, and prior in terms of causality to the outcome measures employed to train the models included in the risk assessment tool.

The latter is a subtle but important conceptual point which addresses two issues: Firstly, by selecting an evaluation measure which is not causally dependent on the outcomes used to train the models, the possibility that assignment to the intervention or control arm in and of itself affects the evaluation is minimized. Secondly, it addresses issues relating to AI system alignment. As suggested by the orthogonality thesis, the predictive performance of an AI system is thought to be independent of the goals of the system as a whole (20). Operationalizing the need for a rapid ambulance response in terms of measurable outcomes is difficult, and we cannot assume that we have done so perfectly. Thus, it is appropriate that both the model and human decisions are evaluated in terms of a measure which in causally independent of either. In this way, the ML framework is not given an unfair advantage over human dispatchers who may have internalized a different definition of patent risk and ambulance care need.

The first secondary outcome in the study is based on a composite score consisting of each of the four outcomes included in the risk assessment instrument. While this measure of intervention effectiveness suffers from the problems noted above, this is the manner in which predictive models are typically evaluated (4,5). The second secondary hypothesis consists of an alternate specification regarding the difference-in-difference of NEWS between prioritized and non-prioritized patients across treatment arms.

### Sample Size

Sample size was determined based on pilot study data indicating that available ambulances were directed to the patient with the highest NEWS 65.3% of the time (considering ties as “correct” assessments), while simulation using randomly selected pairs of potentially eligible patients suggested that the model would mark the patient with the highest NEWS correctly in 70.3% of cases. Using this effect size, an estimated power of 0.8 with an alpha of 0.05 was achieved at n ≈ 1500 using a two-sided test of proportions.

### Randomisation

Patients were recruited by ambulance directors upon identification of an RCS with multiple eligible patients. Study arm allocation was be performed automatically by the server used to generate risk assessments using a simple random number generator implemented by the numpy python package (21).

### Blinding

Patients and the ambulance / hospital staff collecting outcome data were blind to treatment arm allocation, but by the nature of the intervention dispatchers were aware of the randomization results. Data analysis scripts evaluating the primary and secondary hypotheses were written prior to extracting outcome data using synthetic data simulating the null hypotheses. Where it was necessary to manually extract outcome data manually (hospital vital sign data at the second study site), the abstractor was blind to treatment group assignment.

### Statistical analysis

To generate risk predictions, gradient boosting models were applied to patient demographics, structured CDSS data, and free-text notes embedded using the bag-of-words method as described elsewhere (1). The models were implemented in the openTriage platform and accessed by the dispatching system via API (2).

The primary hypotheses was evaluated using logistic regression, with missing vital sign data necessary to calculate NEWS multiply imputed using multivariate imputation by chained equations using the random forest algorithm (22). 10 sets of imputed NEWS components were generated, and full scores were directly calculated from the component items in line with recommendations (23). Per the study protocol, outcomes were to be evaluated based on the median value of 5 imputations, but upon further consideration this was felt to risk inflating type-I error rates, and the more rigorous approach of pooling estimates using Rubin’s rules was used (24). Secondary hypothesis 1 was similarly evaluated using logistic regression, but the second study center had to be excluded due to technical difficulties in gathering comprehensive hospital outcome data. Secondary hypothesis 2 was evaluated using a Wilcoxon rank sum test applied to the multiply imputed data, pooling z-values using Rubin’s rules to test for significance (25).

Six ancillary analyses were pre-specified. Analyses 1-2 investigated compliance with the instrument, and were investigated by assessing the compliance of dispatchers with the risk assessment tool overall and in the high and low confidence groups of the intervention arm, with the hypothesis that compliance would be higher in the high-confidence group. An analysis was also performed to investigate the hypothetical outcomes if dispatchers had been 100% compliant with the tool to evaluate the potential impact of compliance rates. Pre-specified analyses 3-5 regarded changes over time, and were examined using time-series analysis within a regression framework employing a variable representing the study month (or months since last model update) as the independent variable of interest. Analysis 6 regarded model calibration, and was conducted by including patient characteristics of interest (age, sex, and major complaint groups) as independent predictors in models evaluating the primary hypothesis. An analysis of loss to follow-up (i.e., patients who withdrew from the study) was performed to examine whether patients who opted out of the study differed from those to remained. An analysis of time to dispatch was not pre-specified, but was identified as an important indicator of operational efficiency in discussions with the study with clinical and administrative staff.

It was anticipated that dispatchers could attempt to circumvent the randomization process (e.g, repeat the randomization if the RCS was assigned to the control arm), and code was implemented to assign repeated randomizations of the same patients to the same treatment arm. In these cases, only the last randomization was included in the analysis. In cases where randomization was repeated but additional patients were included and thus not captured by the repeated randomization check, the last randomization was only included if all randomizations had by chance been assigned to the same treatment arm. All other randomizations were excluded as protocol violations.

All data transformation and analysis was performed using R v4.4.2 (26), and the scripts used to perform all transformations and analysis are available in a public repository (27). All code to evaluate the prespecified analyses were written prior to obtaining patient outcome data using on a synthetic dataset simulating the null hypothesis. The analysis script used to generate the results presented in this manuscript and its output may be found as supplement 1 – Analysis notebook.

### Ethics

Ethical approval for the study was sought and granted by the Swedish Ethical Review Authority (Dnr 2020-00187). An exemption from gathering prospective informed consent from patients was granted for the study by the ethics review board. Informed consent materials were instead mailed to study participants retroactively, at which point patients were given the opportunity to withdraw from the study. The study and its protocol was preregistered at ClinicalTrials.gov (ID NCT04757194) on 2021-02-17, (28) and is reported according to the CONSORT guidelines (29).

## Results

### Participant flow

A total of 1845 RCS were included for randomization by dispatchers. 350 randomizations (13.5%) were the result of a protocol violation in which no ambulance was dispatched to any patient included in the comparison prior to the next comparison including the same patients. There was also a single case where no ambulance was assigned to any patient in the RCS included in this category of exclusions. Of the remaining 1495 RCS, 250 (16%) included a patient who upon receiving information about the study, opted to decline participation. Upon applying these exclusions, 1245 RCS remained for analysis per figure 1 below.

### Recruitment

Patients were recruited between 2021-02-01 and 2024-12-01. The trial had to be extended due to a slow rate of inclusion and delays in obtaining data necessary to train the models for the second study site, which began data collection in 2024-06-01. A data monitoring committee was formed to monitor the study and address any reported incidents or patient safety issues, but none were identified. The study was ended prior to having collected the full calculated sample size (1500) for administrative reasons.

### Baseline data

A total of 585 RCS were included in the control arm, and 660 in the intervention arm, corresponding to 1285 and 1479 individual patients, respectively. Patient demographics in terms of age and gender were similar across both treatment arms. Overall dispatch times were similar across both treatment arms, but both dispatch times and times from inclusion to dispatch were shorter for prioritized patients in the intervention group. Note that an analysis of dispatch times was not pre-specified.

**Table 1.** Patient / Dispatch characteristics.

|  | Control | Intervention |
| --- | --- | --- |
| Number of RCS Included | 585 | 660 |
| Number of Patients included | 1285 | 1479 |
| Median Age | 77 (76-78) | 77 (76-78) |
| Percent female | 0.45 (0.43-0.48) | 0.44 (0.41-0.46) |
| Median dispatch time (minutes) | 45.1 (42.8-48.6) | 44.6 (42.2-46.8) |
| Median dispatch time for prioritized patient (minutes) | 32.1 (30.4-35.5) | 29.2 (26.5-31) |
| Median time from inclusion to dispatch for prioritized patient (seconds) | 202 (176.1-264.5) | 95.6 (81.5-135) |

*All values are reported as central tendencies and 95% confidence intervals based* *on percentiles of 1000 ordinary bootstrap samples.*

### Missingness

NEWS score calculation was based primarily on ambulance vital parameters, which were missing in between 11.0% (pulse) and 14.2% (temperature) of cases. Missing ambulance data could be due either to patients not receiving an ambulance, or ambulance staff not fully documenting vital parameters. Ambulance vital sign data was supplemented with hospital data collected from the patient’s first ED visit within 24 hours of contact with the EMD center, resulting in final rates of between 3.7% (consciousness) and 4.8% (temperature). These combined datasets were used to perform multiple imputation which achieved stable imputed values with good mixing properties across chains. Full data on missingness rates and imputation diagnostics may be found in supplementary materials 1.

### Primary and secondary hypotheses

In the control arm, the patient with the highest NEWS value was prioritized in 62.5% of cases, while in the intervention arm the proportion was 68.3%, corresponding to an odds ratio of 1.28 (95% CI 1.00 – 1.63, p = 0.047). There is thus support for rejecting the null regarding the primary hypotheses of the study.

Similarly, intervention effects regarding the first secondary hypotheses achieved statistical significance, with 63.0% of prioritized patients in the control arm having the highest composite outcome score, compared to 69.6% in the intervention arm, corresponding to an odds ratio of OR 1.31 (1.01 – 1.72, p = 0.041) among the 1106 RCS in the first study site. The average difference between the NEWS value of prioritized and non-prioritized patients in the control arm was 0.62, compared to 1.08 in the intervention arm, corresponding to a mean difference of 0.45, although based on a Wilcoxon rank-sum test, no significant difference could be identified (p = 0.086). Results are summarized in table 2 below.

| Hypothesis | Result |
| --- | --- |
| Primary: Odds of prioritized patient having the highest NEWS in an RCS is greater in the intervention arm | OR 1.28 (95% CI 1.00 – 1.63, $p = 0.047$) |
| Secondary: Odds of prioritized patient having the highest composite outcome score in an RCS is greater in the intervention arm | OR 1.31 (1.01 – 1.72, $p = 0.041$) |
| Secondary: Larger difference in NEWS scores between prioritized and non-prioritized patients in intervention arm per a rank sum test | Z-score 1.716, $p = 0.086$ |

### Ancillary analyses

Six additional pre-specified analyses were performed to evaluate the properties of the study and the risk prediction model itself.

Per the study protocol, directors were permitted to deviate from the model recommended patient upon conferring with the involved dispatch nurses. In the control arm where information regarding the risk score was not available, the patient with the highest risk score was prioritized in 54.0% (50.0 – 58.1) of cases. In the intervention arm, the patient with the highest risk score was prioritized in 80.9% (77.7 – 83.9) of cases. To evaluate the impact of these deviations, an analysis was performed to estimate the intervention effect had the compliance with the risk score been 100% in the intervention group. This analysis identified a substantially stronger effect, with 72.2% of prioritized patients having the highest NEWS value, and resulting in an odds ratio of 1.56 (1.21 – 2.00, p < 0.001).

An indicator of model confidence was included in the intervention as described in the methods, and while a difference in compliance between the high- and low-confidence groups was identified (81.7% vs 80.3%), the difference was not found to be significant, nor was there a significant difference in outcomes between the groups as assessed per the primary hypothesis. There was however a substantial difference when analysed in the 100% compliance scenario, with 67.4% of patients assessed by the instrument as highest risk having the highest NEWS value in the low confidence group, vs 78.5% in the high confidence group, corresponding to an OR of 1.77 (1.20 – 2.62, p = 0.004) between the confidence groups.

A number of analyses investigating temporal effects during the study period were also performed. It was hypothesized that compliance with the tool might change over the study period (either positively due to routinization effects, or negatively due to loss of trust in the tool). The per-month change in the odds of compliance with the tool in the intervention group was 0.98 (0.97 – 1.001, p = 0.07).

It was also hypothesized that the dispatchers might learn from the assessments of the tool, resulting spillover to the control group, manifesting as an increased assessment accuracy over time in the control group. No evidence of this could be found however, with a per month change in the odds of correct assessment per the primary hypothesis in the control group of 1.00 (0.98 – 1.01, p = 0.944).

There was also concern that model performance could degrade over time, and model performance was monitored over the course of the study by the Data Monitoring Committee in terms of the correlation between the patients assigned risk score and their resultant NEWS value. A risk of degradation was thought to have been identified once early in the study, and the model was retrained with updated data. The model was updated once more during the course of the study upon initiation of the study at the second site. The updates were made on the first of December 2021 and the first of June 2024, respectively. Upon analysis of the full dataset however, no linear degradation of model performance could be identified, with an average change in spearman correlation per month since the most recent update of -0.00 (-0.01 – 0.01, p = 0.906).

To evaluate model calibration, indicators for patient age, sex, and clinical category were used as predictors of NEWS together with the ML risk score. We found no residual predictive value of patient age or sex in predicting NEWS when adjusted by the ML risk score. Significant predictive values for patient categories were identified for 2 of the 41 patient types included in the study (Fever and Difficulty breathing). Full results regarding all ancillary analyses may be found in the supplementary materials.

## Discussion

This study evaluated the ability of a ML-based risk scoring tool to influence care providers at EMD centers, with the aim of improving prioritization decisions of low priority cases in resource constrained situations. The intervention was found to have resulted in an improved differentiation of patients, though intervention effects were at the edge of statistical significance. The intervention appeared to be stable over the nearly 4-year study timeframe, with no signs of degraded model performance. In addition to improved differentiation, the intervention group also had shorter dispatch delays for prioritized patients. While this analysis was not prespecified, these findings suggest that the intervention may also have improved the speed of the dispatching process.

This is, to our knowledge, the first randomized trial of a ML-based risk assessment tool intended for use in the general patient population served by EMD centers. In a previous RCT intended for use in identifying cardiac arrests at EMD centers, Blomberg et al. (10) identified no intervention effect of an ML-based alerting tool, which was attributed to a lack of compliance with the intervention by EMD nurses. These findings, along with the results of our study identifying the potential for a substantially larger intervention effect had the tool been followed more closely, highlight the need to understand how to build trust in automated risk assessment tools if they are to be used to their full potential. An indicator of model confidence was included in our intervention, but it failed to achieve a statistically significant impact on compliance, despite the high-confidence intervention arm RCS containing a higher proportion of accurate assessments. While the definition of the outcome metrics employed was largely based on clinical judgement, the similar effect sizes with regards to predicting NEWS (OR 1.28) and the composite outcome the model is trained to predict (OR 1.31) suggests that the risk predictions were well aligned with a widely used risk differentiation tool.

While our study identified a statistically significant intervention effect, the overall level of accuracy leaves much to be desired, highlighting the difficulty of triaging potentially emergent conditions over the telephone. In order to improve the accuracy of the ML models used, we see two general paths: The inclusion of unstructured audio data from the call, and the inclusion of additional structured data from historical patient medical records. The former may be accomplished e.g. through the processing of audio data from the emergency call through pretrained speech recognition models, and its integration with the decision support system data. The challenges of integrating patient medical record data are primarily legal and technical, necessitating the availability of APIs for obtaining high-quality data from medical records systems in near real-time. Nonetheless, our findings suggest that the effect size hypothesized based on retrospective validation indeed translated into a real-world impact of similar magnitude, and that applications aimed at higher-risk patient groups using risk assessment tools with similar levels of performance may be safely pursued.

### Limitations

By the nature of the intervention, dispatchers could not be blinded to the treatment assignment. This could explain the uneven distribution between the intervention and control arms (660 vs 585), as dispatchers sometimes sought to repeat the risk scoring if they were assigned to the control group, resulting in these repeat risk assessments sometimes being excluded due to protocol violation. While this exclusion mechanism does not appear likely to bias the results, it reduced the sample size and thus the power of the analysis. Missing outcome data also reduced the power of the analysis by inducing between-imputation variability, resulting in an effect size similar to that expected based on pilot study results, but with a substantial degree of uncertainty. The multiple imputation process is also stochastic, and given that the findings are at the very edge of statistical significance, even simply changing the random seed used to generate imputations can variously produce significant or non-significant findings. We thus urge that the p-values reported here be interpreted thoughtfully as the probability of repeated trials generating an effect at least this extreme under the null hypothesis, rather than dichotomously.

This study was performed at dispatch centres employing nurses in the primary call-taking role. This level of formal education is relatively rare in the context of EMD and could impact the generalizability of the results. It is reasonable to believe that it would be in the direction of underestimating the intervention effects were models of similar precision implemented in a context where the control group were assessed by care providers with less formal training. Similarly, the models used are based on a CDSS used only in a small number of Swedish regions. However, the modelling framework is freely available and can be adapted to structured and free-text data from other CDSS.

The study suffered from a degree of post-randomization drop-out due to the 306 RCS (16%) having to be excluded due to at least one patient opting out of the study. The characteristics of patients excluded due to this were however similar to those included in the study (see supplementary materials 1), and this source of loss to follow-up thus does not appear to have impacted the findings.

## Conclusion

This randomized controlled trial suggests that ML-based interventions in the context of emergency medical dispatching have the ability to improve the capacity of care providers to identify patients most in need of an ambulance across a diverse patient cohort. The overall accuracy of the triage process however remains modest, and more can be done to improve the accuracy of the models and the adherence of care providers with model recommendations.

## Data Availability

The data used in this study are owned by the regional health authorities in each respective study site, and permission to publicly distribute individual level data was not granted by the Swedish ethics review authority. The data used in this study may be obtained by researchers with appropriate ethics approvals by contacting. All code used to generate the reported results are available in a public repository at https://osf.io/erkv7/ and the tool evaluated is available at https://github.com/dnspangler/opentriage

## Funding

Partly funded by the Swedish Innovation Agency grant number 2017-04652. The funders had no role in study design, data collection and analysis, decision to publish, or preparation of the manuscript.

## Conflicts of interest

The authors have declared that no competing interests exist.

## Patient or Public Involvement

No formal Patient or Public Involvement efforts were involved in this study.

## Notes

### Competing Interest Statement

The authors have declared no competing interest.

### Clinical Trial

ClinicalTrials.gov ID NCT04757194

### Clinical Protocols

https://clinicaltrials.gov/study/NCT04757194

### Funding Statement

Yes

### Author Declarations

Swedish Ethical Review Authority Dnr 2020-00187

